# Exploring Patient Centered Perspectives on Suicidal Ideation: A Mixed Methods Investigation in Gastrointestinal Cancer Care

**DOI:** 10.1101/2023.12.11.23299772

**Authors:** Avishek Choudhury, Yeganeh Shahsavar, Imtiaz Ahmed, M Abdullah Al-Mamun, Safa Elkefi

## Abstract

**Background:** Gastrointestinal (GI) cancer patients face a 4-fold higher risk of suicide than the general US population. While efforts to reduce suicide risk in cancer patients have been crucial, the issue remains urgent. This study explores the psychosocial aspects of gastrointestinal (GI) cancer patient experiences. It focuses on assessing the prevalence and intensity of suicidal ideation and behavior, evaluating mental distress during different treatment phases, and examining the impact of psychosocial factors on mental health.

**Method:** We used a two-phase mixed-methods approach. The research began with a web-based survey targeting individuals across the US who were undergoing or had completed GI cancer treatment. Survey respondents were then debriefed and invited to engage in a follow-up interview, which were conducted via video conferencing, recorded, transcribed, and anonymized for analysis. Quantitative data analysis involved confirmatory factor analysis to validate constructs related to mental health and suicidal ideation. Subsequently, significant correlations with mental health and suicidal ideation were extracted and scaled for further bivariate analyses. Qualitatively, an inductive thematic analysis was performed on both survey responses and interview transcripts. The patient journey was charted from diagnosis through the treatment continuum, with sentiment analysis enriching the thematic findings.

**Results:** Two hundred-two individuals responded to the survey. Of all the participants, 76 respondents were from the rural Appalachian region, and 78 were undergoing treatment during the study. Quantitative analysis revealed a higher prevalence of passive suicidal ideation compared to active planning. The post-treatment recovery period was reported as the most emotionally challenging. Qualitative data emphasized the crucial role of emotional support, with patients feeling particularly vulnerable to isolation. The quality of care received was another concern, with calls for more individualized treatment plans and better communication. Patients also expressed a need for clear, comprehensive information about their treatment and potential side effects. The in-depth interview with 4 GI cancer patients indicated a healthcare system that prioritizes expedient treatment over comprehensive care, with a noted lack of formal psychological support. The role of artificial intelligence (AI) emerged as a promising avenue for enhancing patient understanding of their condition and treatment options. Patients valued clear, empathetic communication and the provision of comprehensive, understandable information from their healthcare providers. The sentiment analysis associated with their experiences reflected a spectrum of emotional responses, from shock and disbelief at diagnosis to fluctuating emotions during treatment.

**Conclusion:** Our research advocates for a more patient-centric model of care, enhanced by the thoughtful integration of technology and consistent, empathetic communication. These findings contribute to a deeper understanding of the GI cancer patient experience and provide a foundation for improving cancer care practices to better address the holistic needs of patients.

## Introduction

Suicide constitutes a pressing global public health issue, exerting a substantial impact on mortality rates across the world [1]. The World Health Organization (WHO) reports an estimated 703,000 annual suicide-related deaths worldwide. Moreover, incidents associated with suicide, such as contemplation of suicide, suicide attempts, devising suicide plans, and impulsive thoughts of self-harm, can inflict adverse psychological consequences on both individuals and those in their immediate social circles [2].

According to the WHO’s 2019 assessments, cancer emerges as one of the leading cause of premature death before the age of 70 across 112 nations [3]. One of the most disturbing factors linked with cancer is the significantly increased risk of suicide among patients. This issue has attracted considerable attention from the medical community and society [4–11]. Studies reveal that gastrointestinal (GI) cancer, a group of cancers that affect the gastrointestinal tract and associated organs (digestive system), patients face a 4-fold higher risk of suicide than the general US population, with suicide ideation and completion being most prevalent within the first six to twelve months post diagnosis [7,12,13]. This disturbing trend has prompted several health organizations to take notice. The American College of Surgeons Committee on Cancer, the American Society of Clinical Oncology, and the National Comprehensive Cancer Network have all stressed the importance of addressing cancer patients’ psychological and emotional needs [14,15]. While efforts to reduce suicide risk in cancer patients have been crucial, the issue remains urgent.

The psychological impact of GI cancer, including depressive disorders and suicidal ideation, has been well-documented, with studies highlighting the strong relationship between psychological characteristics and internet use, family burden, household occupancy, age, and emotional support [16]. Furthermore, coping style, insomnia, and psychological distress have been identified as important factors affecting the well-being of individuals with gastrointestinal cancer, emphasizing the need for a holistic approach to address the psychological aspects of this patient population [17]. The prevalence of suicidal ideation in cancer patients, including those with gastrointestinal cancers, has been a subject of research, with studies reporting a higher risk of suicidal ideation in specific cancer subtypes, such as ovarian cancer [18]. The distress experienced by cancer patients, as measured by tools like the Distress Thermometer, has been found to have predictive value for assessing the risk of suicide in this population, highlighting the need for proactive psychological support and intervention [19]. In cancer care, acknowledging and incorporating the voices, perspectives, and needs of GI cancer patients is increasingly recognized as a critical aspect of comprehensive healthcare. This perspective emphasizes the necessity of understanding and addressing the unique challenges faced by individuals with GI cancer, aiming to enhance both their treatment outcomes and overall quality of life.

As healthcare providers and researchers strive to optimize care, the importance of patient-centered approaches becomes self-evident. In this study, we explore the needs and stories of GI cancer patients, emphasizing the potential impact of personalized, holistic care on suicidal ideation and mental distress in affected individuals. Our investigation seeks to explore patient perspectives, offering insights that could reshape the landscape of cancer care. By doing so, we aim to foster a more compassionate, comprehensive, and practical approach to addressing the psychological challenges often accompanying a GI cancer diagnosis.

## Methods

Our study received ethical approval from the West Virginia University Institutional Review Board under protocol number 2212691613. The study qualified for the WVU Flexibility Review Model, as it involves minimal risk and adheres to the Belmont Report’s ethical principles. Approval was granted on February 7, 2023.

As illustrated in Figure 1, the data for this study was collected in two sequential phases (A and B) through a validated web-based survey and semi-structured interviews, respectively. Any individual in the US with GI cancer (undergoing or completed treatment) was eligible to participate in this study.

**Figure 1.**
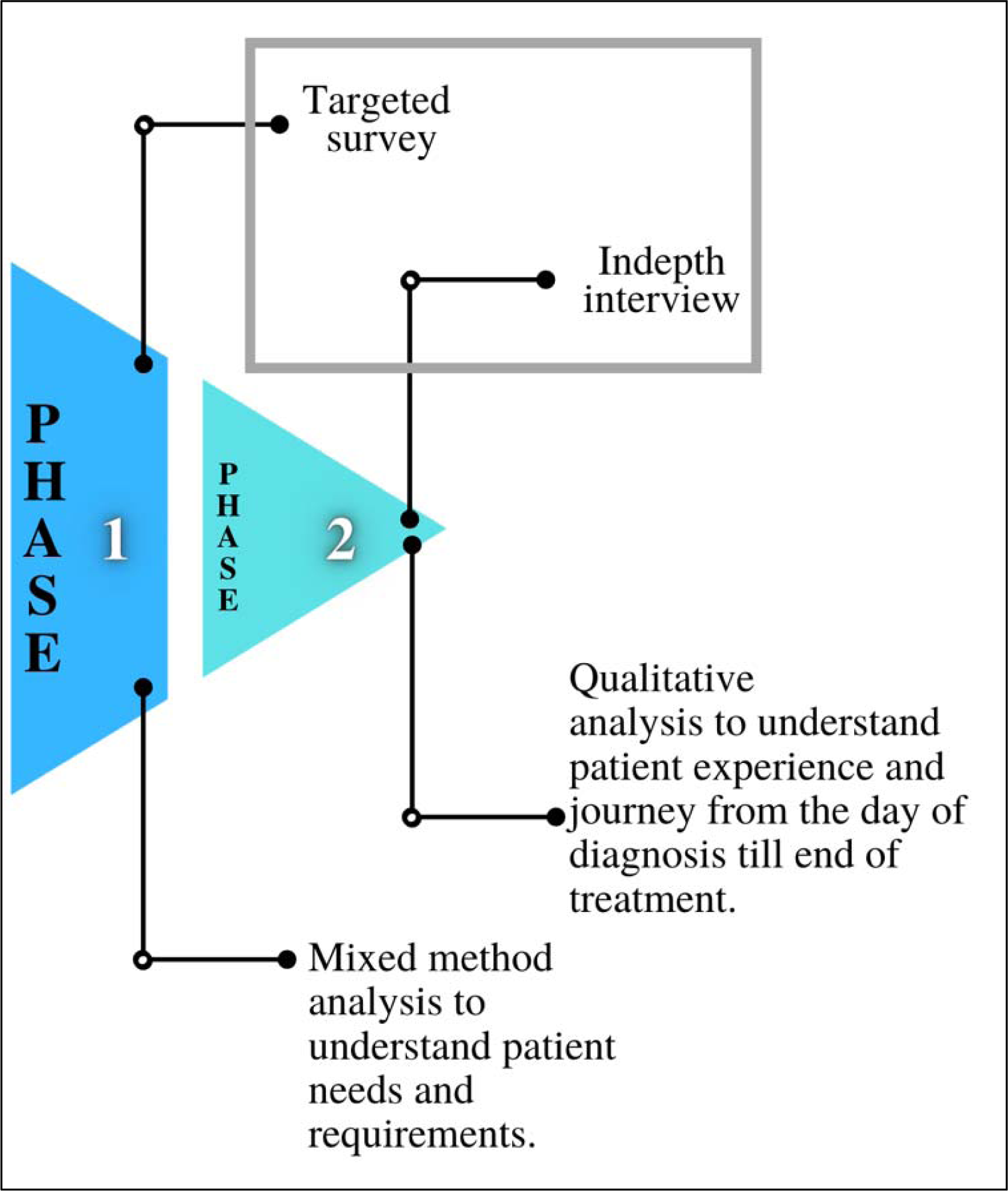
The two phases of study methodology

In phase 1, a web-based survey (see Figure 2) was distributed to cancer patients through social media and open cancer forums via an audience paneling service, Centiment. Centiment reaches a broader and more representative audience via its network and social media. They also use fingerprinting technology that combines IP address, device type, screen size, and cookies to ensure that unique panelists enter the survey.

**Figure 2.**
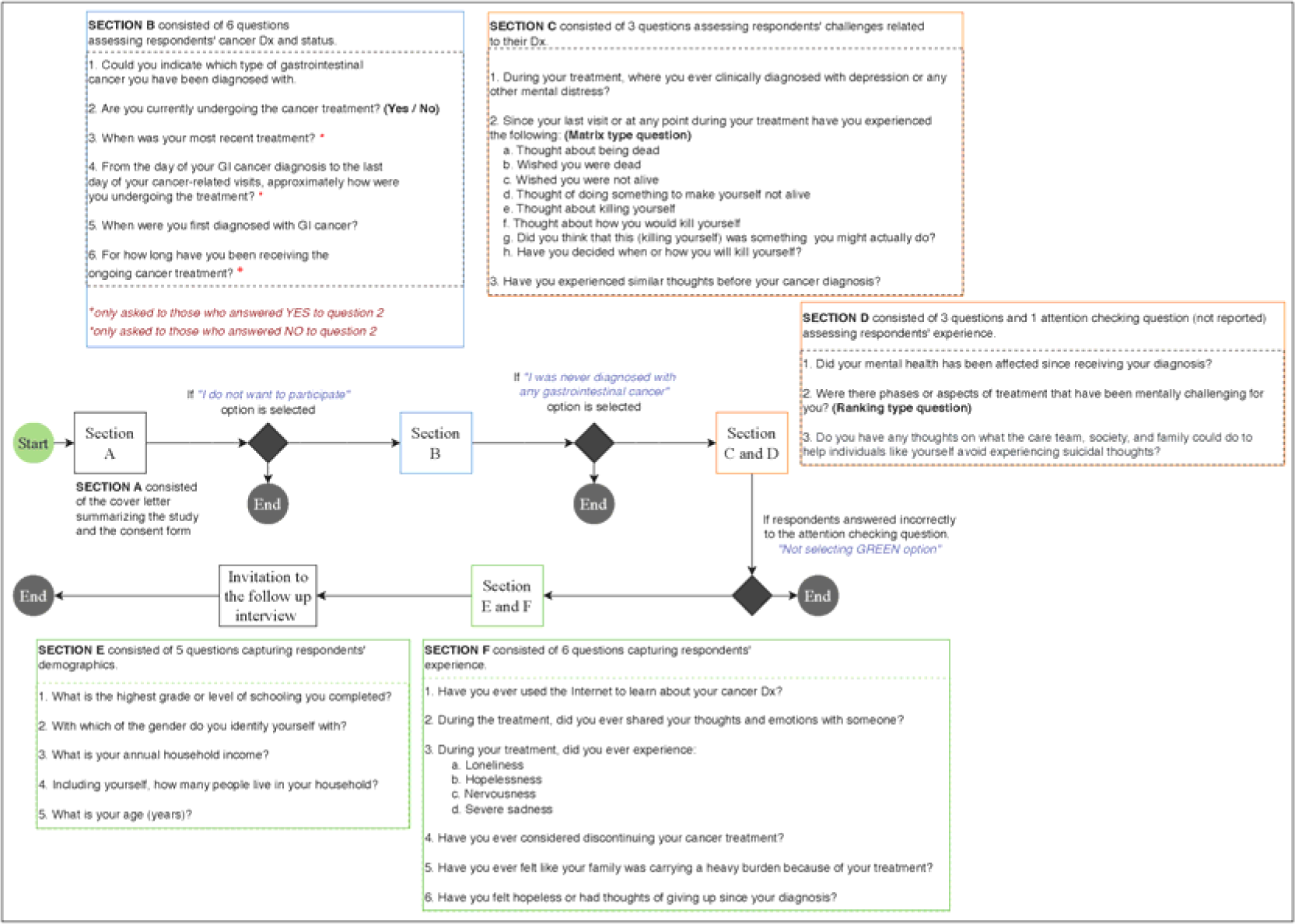
Survey design and flow

The survey contained validated questions adapted from the Columbia-Suicide Severity Rating Scale (C-SSRS) [20] and the Patient Health Questionnaire (PHQ-4) [21,22]. Other variables, such as family burden, internet information utilization, and emotional support, were measured using a five-point Likert scale.

The survey was accompanied by a detailed cover letter, an electronic consent form, and an invitation for an online follow-up interview. After reviewing the cover letter and consent form, individuals with gastrointestinal issues who agreed to participate were directed to the survey. Participant eligibility was determined through self-reported data. A control question was included to maintain response integrity: “To confirm attentive reading and thoughtful responses, please select Green as your answer.” Responses failing this criterion were excluded from the analysis.

Participants were debriefed and invited for a follow-up interview after completing the survey. The principal investigator’s (PI’s) contact information was provided, encouraging participants to reach out if interested in the interview phase. Four individuals contacted the PI and agreed to participate in the follow-up interview (i.e., Phase 2).

Phase 2 started soon after the completion of Phase 1. The PI interviewed the participants over Zoom for approximately 45 minutes per participant. Both the parties had their web camera switched on. The interview sessions were recorded and manually transcribed for analysis. After each interview, the participants were emailed a $50 Amazon gift card as an honorary. No personal identifiers were collected. All names and identifiers in the transcription were anonymized before analysis. Figure 3 shows the overall interview process.

**Figure 3.**
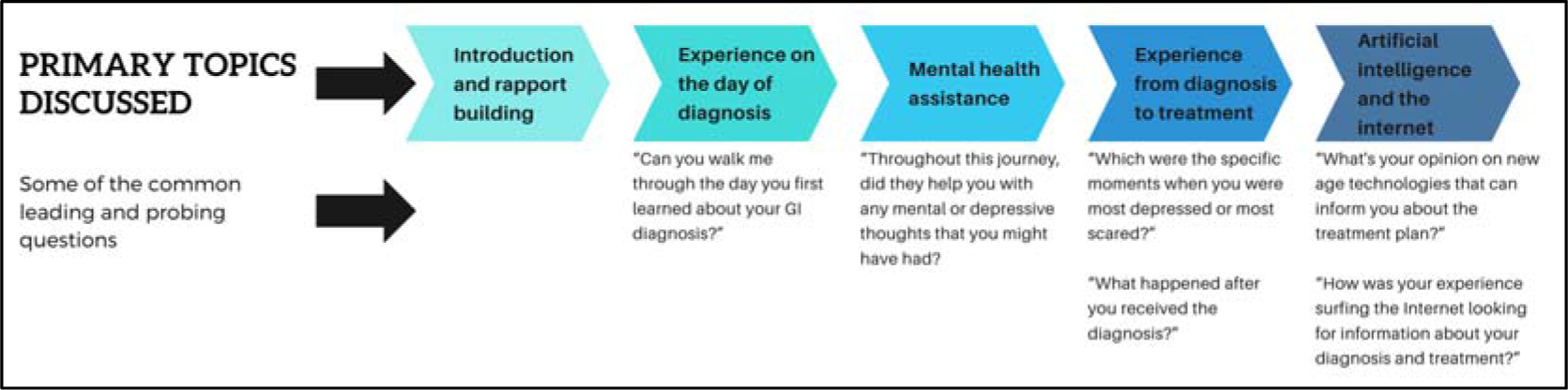
The overall interview processes.

### Quantitative analysis

The descriptive statistics of all the variables were calculated. The questions adapted from PHQ-4 and C-SSRS were converted into latent constructs, namely mental health (Factor 1) and suicidal ideation (Factor 2), respectively. The constructs were validated using multigroup confirmatory factor analysis across both groups (participants from rural Appalachia and others). Model fit was assessed by the comparative fit index (CFI) [23], Tucker Lewis Index (TLI) [24], and Bentler Bonnet Normed fit index (NFI) [23].

Additional measures such as the parsimony normed fit index (PNFI) [25], Bollen’s relative fit index (RFI) [26], Bollen’s incremental fit index (IFI) [26], and relative non-centrality index (RNI) [27] were also observed. Reliability and validity were assessed using Cronbach’s alpha requiring to be greater than 0.70; (b) outer loadings greater than 0.50 [28] Fornell-Larcker criterion [29] and the Heterotrait-monotrait (HTMT) ratio required to be less than 0.85 [30]. The reliability and validity outcomes are reported in ***Appendix A***.

Upon validating the latent constructs, the scores of each construct were extracted, and the dataset was scaled before a Bayesian Pearson’s Rho correlation was conducted. Next, the significant correlations with mental health and suicidal ideation were identified, and pairwise bivariate analyses were conducted to observe further insights.

### Qualitative thematic analysis

An inductive thematic analysis was employed to analyze the open-ended survey responses and the in-depth interview script. Three coders (two with experience in qualitative data analysis and one with expertise in the subject matter) reviewed each response. Upon discussing with each other, they assigned tentative labels (or codes) in Microsoft Excel, [31] a process called open coding. [32] The coders then reviewed the responses and open codes to identify relationships and similarities between codes, a process known as axial coding. [33] The codes were merged into broader themes representing the core concepts, called selective coding. [34] All coders engaged in iterative discussions to refine and finalize a set of consensus codes, ensuring that the identified themes accurately captured the essence of the participants’ experiences and perspectives.

Additionally, the in-depth interview transcripts were analyzed to chart the patient’s experience, beginning with the day they received a cancer diagnosis and continuing through the entire treatment and recovery process. First, three coders iteratively read the transcripts to understand participants’ perceptions of their journey better. Then, with discussion and consensus, the team of coders matched the participants’ responses with the phase in their cancer journey (day of diagnosis and during treatment).

The organized responses were analyzed using the syuzhet package in R [35]. This package is designed for sentiment analysis, drawing upon a variety of sentiment dictionaries to evaluate the emotional valence of text [35]. This sentiment was subsequently interpreted in conjunction with the findings from the thematic analysis.

## Results

### Participant characteristics

Two hundred and two individuals participated in the study. Of all the participants, 76 respondents were from the rural Appalachian region, and 78 were undergoing treatment during the study. About 73 respondents were not clinically diagnosed with mental distress during treatment. The demographic trends indicative of the diversity within the patient population emerged. Educational level varied across the cohort, with a substantial number achieving some college-level education. Notably, 54 individuals reported some college education without obtaining a degree, while 50 completed a 4-year degree. Those with professional degrees numbered 25, and 6 individuals had attained a doctorate level of education. Income levels demonstrated an economic spread across the cohort, with a significant number of participants (44) reporting annual household incomes between $20,000 to <$35,000, followed closely by those earning $50,000 to <$80,000 (42 individuals). Fewer participants reported incomes at the extremes, with 23 individuals earning less than $20,000 annually and 7 earning more than $200,000, underscoring the economic diversity within the study group. Age demographics highlighted a concentration of older adults, with 72 participants older than 65, the largest group in the cohort. The remaining age groups were more evenly distributed, with the 46 to 55 years and 56 to 65 years categories being the next most populous, with 54 and 41 individuals, respectively. Notably, the younger age groups (18 to 24 and 25 to 35) were represented by only one individual each, indicating a predominance of middle-aged to older adults in the study. Figure 4 illustrates the response flow among a set of survey questions.

**Figure 4.**
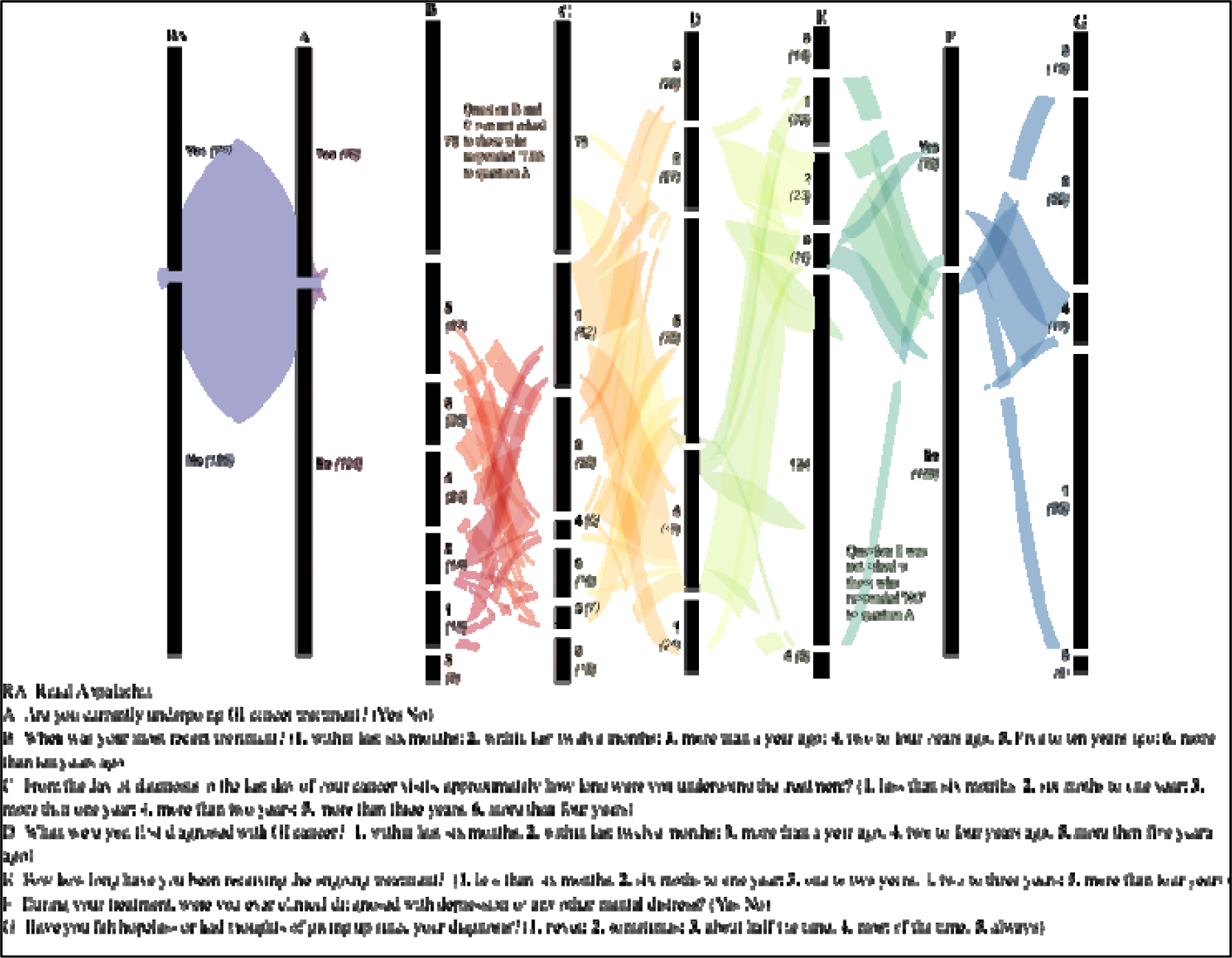
Sankey diagram illustrates the flow and distribution of participant responses through a set of sequential surveys with conditional branching. Each vertical band represents a different question from the survey, with the width of the bands and connecting ribbons proportional to the number of responses. The leftmost band (RA) bifurcates respondents into two categories, ‘from rural Appalachia (’Yes’ 76) or not (’No’ 126), with subsequent questions contingent upon prior answers. The thickness of each flow represents the number of respondents transitioning from one question to the next.

### Quantitative findings

Figure 5 demonstrates the prevalence of suicidal ideation and behaviour among study participants (GI cancer patients). Initial questions regarding passive suicidal ideation, such as the desire to be dead or not wake up, elicited a higher number of affirmative responses compared to more active forms of ideation, such as having a suicide plan. The data reveal an intriguing decline in affirmative responses as the questions probe more concrete steps toward suicidal action. For example, while 78 individuals contemplated death, only 31 had progressed to formulating a specific plan. This decrement underscores the potential for intervention strategies to disrupt the progression from ideation to action.

**Figure 5.**
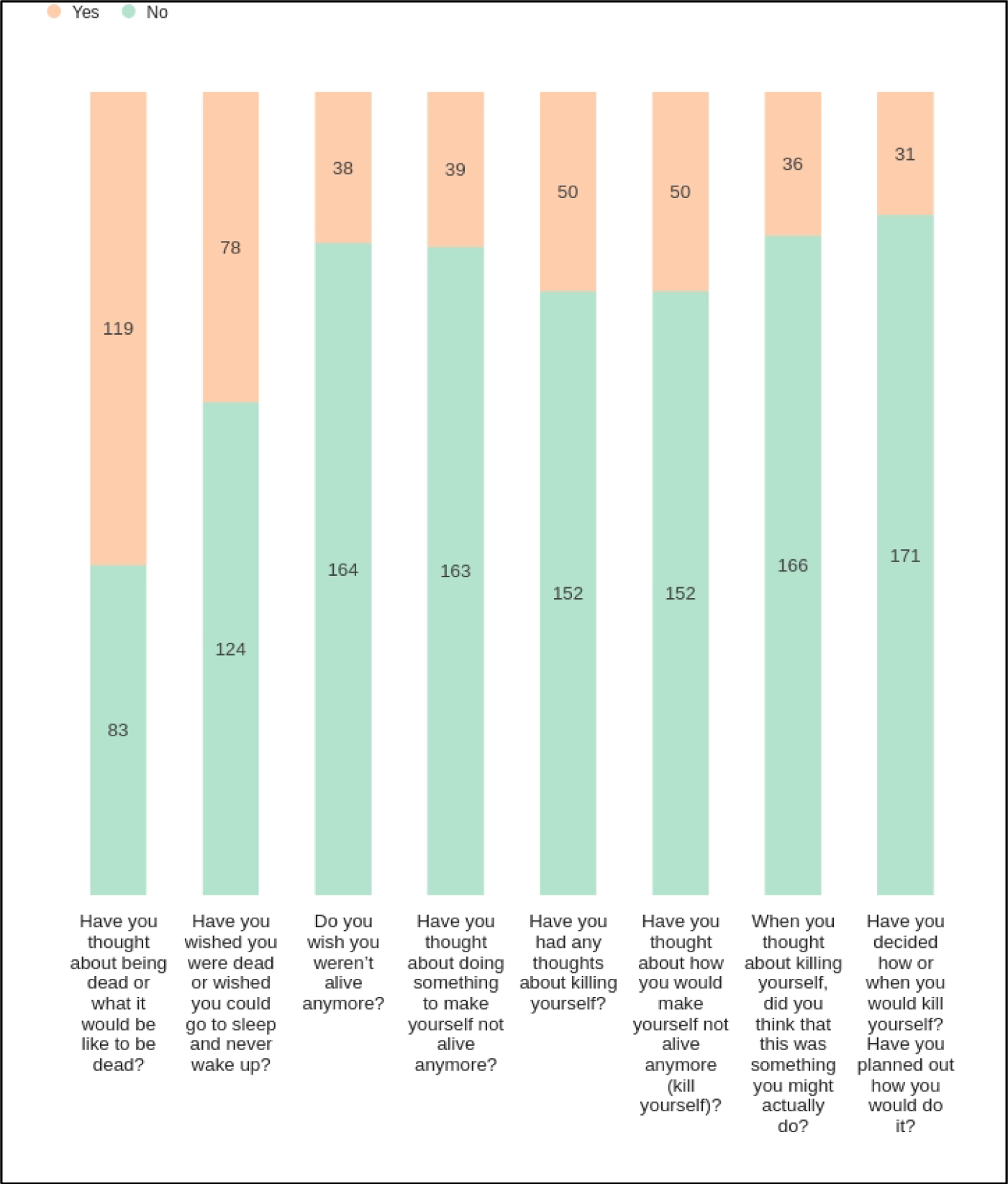
Distribution of Responses to Questions on Suicidality. The figure presents a stacked bar chart depicting participant responses to a series of questions related to suicidality. Each bar represents the number of ‘Yes’ and ‘No’ responses to individual questions, with the total height corresponding to the total number of respondents. The questions progress from less severe ideation (e.g., thinking about being dead) to more severe considerations (e.g., planning how to kill oneself). The colors differentiate between ‘Yes’ responses (peach) and ‘No’ (teal), with the respective counts provided within the bars.

Figure 6 presents a histogram, with each bar representing the number of cancer patients who ranked particular phases of their treatment journey according to the level of mental distress experienced, from 1 (least distressing) to 5 (most distressing). It indicates that patients may find the recovery period after the end of treatment to be the most challenging in terms of mental and emotional health, which could inform healthcare providers about when to offer increased mental health support. Conversely, the day of diagnosis, which might be assumed to be highly distressing, appears to be less so for the respondents in this survey. This could reflect various factors, including relief at having a precise diagnosis or effective initial counseling and support.

**Figure 6.**
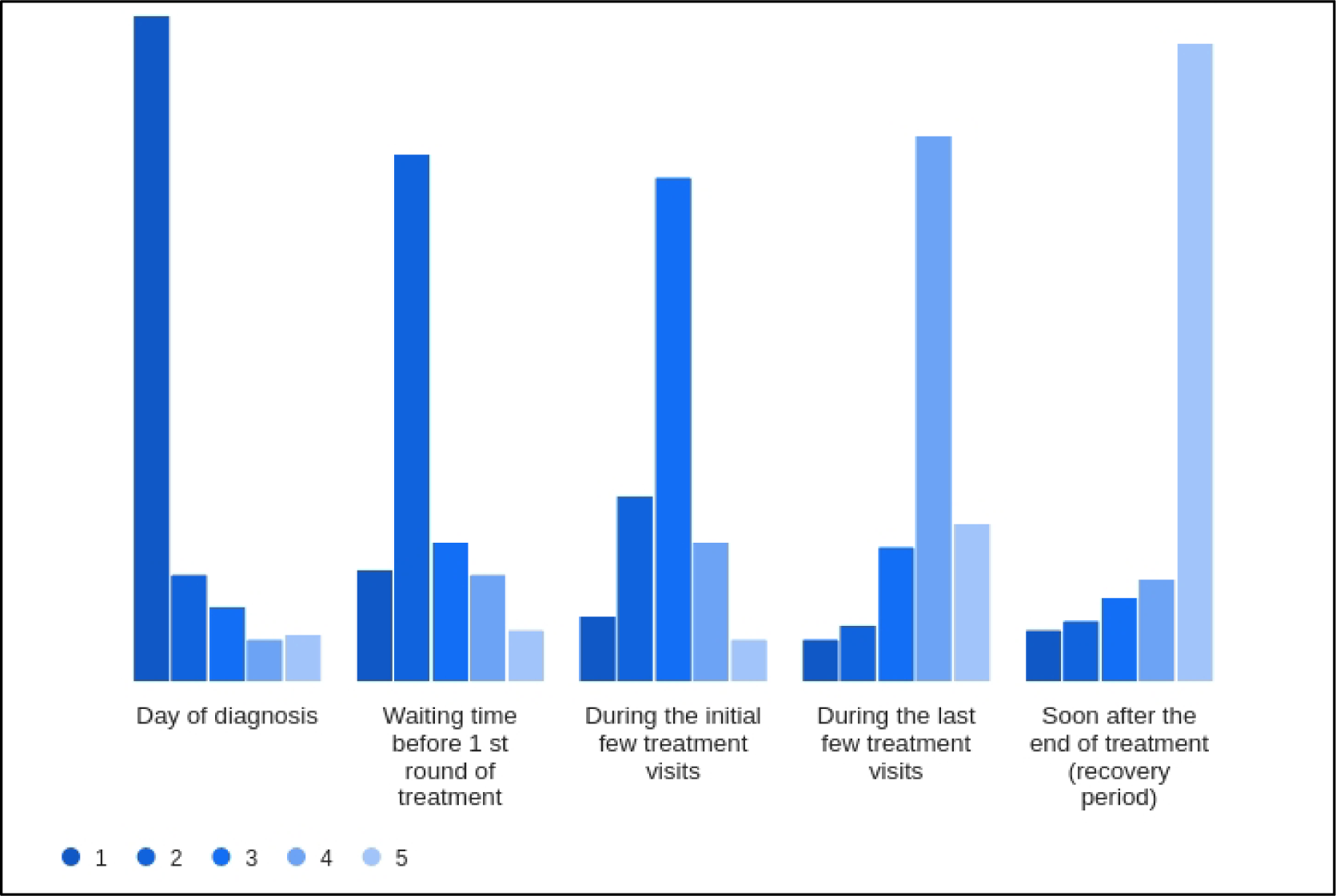
Patient-Reported Mental Distress During Different Phases of Cancer Treatment. The histogram illustrates the distribution of mental distress rankings assigned by 202 cancer patients to five distinct phases of their cancer treatment journey. Each phase is evaluated on a scale from 1 (least distressing) to 5 (most distressing). The results show that the ‘Day of diagnosis’ is most frequently ranked as least distressing, whereas the ‘Soon after the end of treatment (recovery period)’ is predominantly ranked as the most distressing phase. This counterintuitive finding highlights patients’ complex emotional landscape and underscores the need for supportive care throughout the treatment continuum, especially post-treatment.

Table 1 presents descriptive statistics for study variables. The analysis of the mean scores for suicidal ideation (SI 1-8) items reveals significant insights into the prevalence (as shown in Figure 5) and intensity (mean reported in Table 1) of these thoughts among the surveyed individuals. For instance, when both the mean SI scores (Table 1) and the number of ‘Yes’ responses in the corresponding Figure 5 are high, it suggests that a specific aspect of suicidal ideation is commonly experienced among respondents. This convergence indicates a shared tendency toward such thoughts, pointing to a generalized concern that may warrant public health attention. Conversely, lower mean SI scores (Table 1), when juxtaposed with a substantial number of ‘Yes’ responses (Figure 5), suggest a more polarized experience within the population. This dichotomy implies that while the average level of suicidal ideation may be moderate, there exists a subset of individuals experiencing intense levels of such ideation, thereby revealing a hidden severity not immediately apparent in the mean score alone.

**Table 1.**
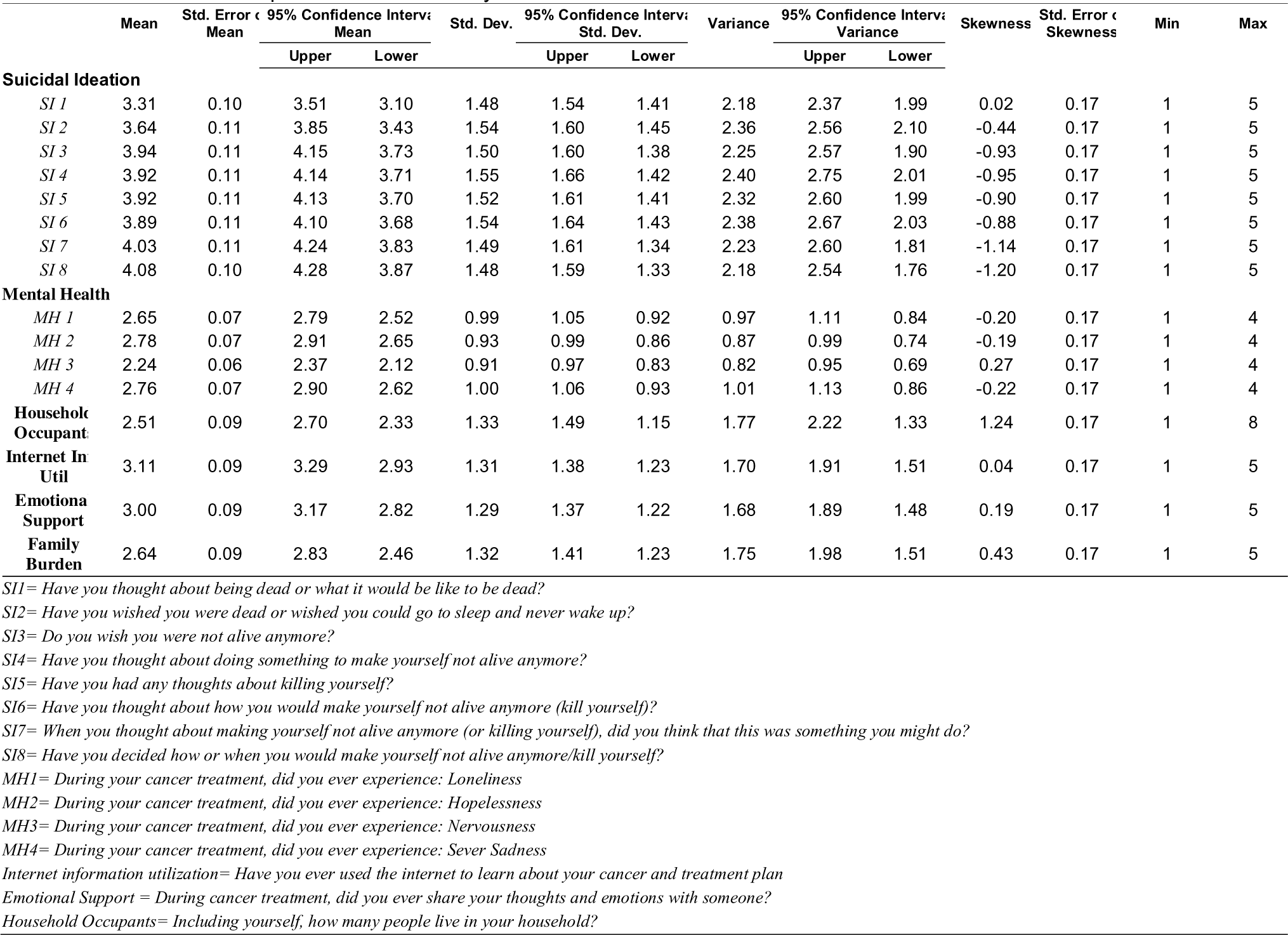
Descriptive statistics of study variables.

Furthermore, the standard deviations (Std. Dev.) and variances suggest moderate response variability. Notably, the 95% confidence intervals for mean scores of SI items remained narrow, reflecting a high level of precision in the measurement of central tendency. However, a negative skewness in the latter items (SI 3-8) suggests a tendency towards lower frequencies of more severe suicidal ideation, with fewer respondents contemplating concrete plans or actions.

The mental health experiences depicted by MH 1-4 (Table 1) show a similar pattern of narrow confidence intervals and low skewness, with mean scores indicating less distress than the SI items. However, the negative skewness in three out of four MH items suggests that a significant number of individuals are experiencing less favorable mental health states, which the averaging effect of the mean could mask.

The utilization of internet information (Internet Info Util) and seeking Emotional Support have mean scores around the mid-range of the scale, suggesting a moderate engagement with these resources among respondents. The skewness in the Family Burden item is notably positive, indicating that many respondents perceive their family as facing a substantial burden due to their treatment.

In the examined cohort of cancer patients, our correlation analysis (Figure 7) delineated the relationship between several psychosocial factors and both mental health and suicidal ideation. A significant inverse correlation was found between mental health and suicidal ideation (r = -0.434, *p < .001*). Access to information and emotional support also played important roles. Internet information utilization was inversely related to suicidal ideation (r = -0.280), proposing that the ability to find health-related information online might serve as a protective factor against the development of suicidal thoughts. In contrast, emotional support was positively correlated with better mental health (r = 0.274), underscoring its importance in the psychosocial health of cancer patients.

**Figure 7.**
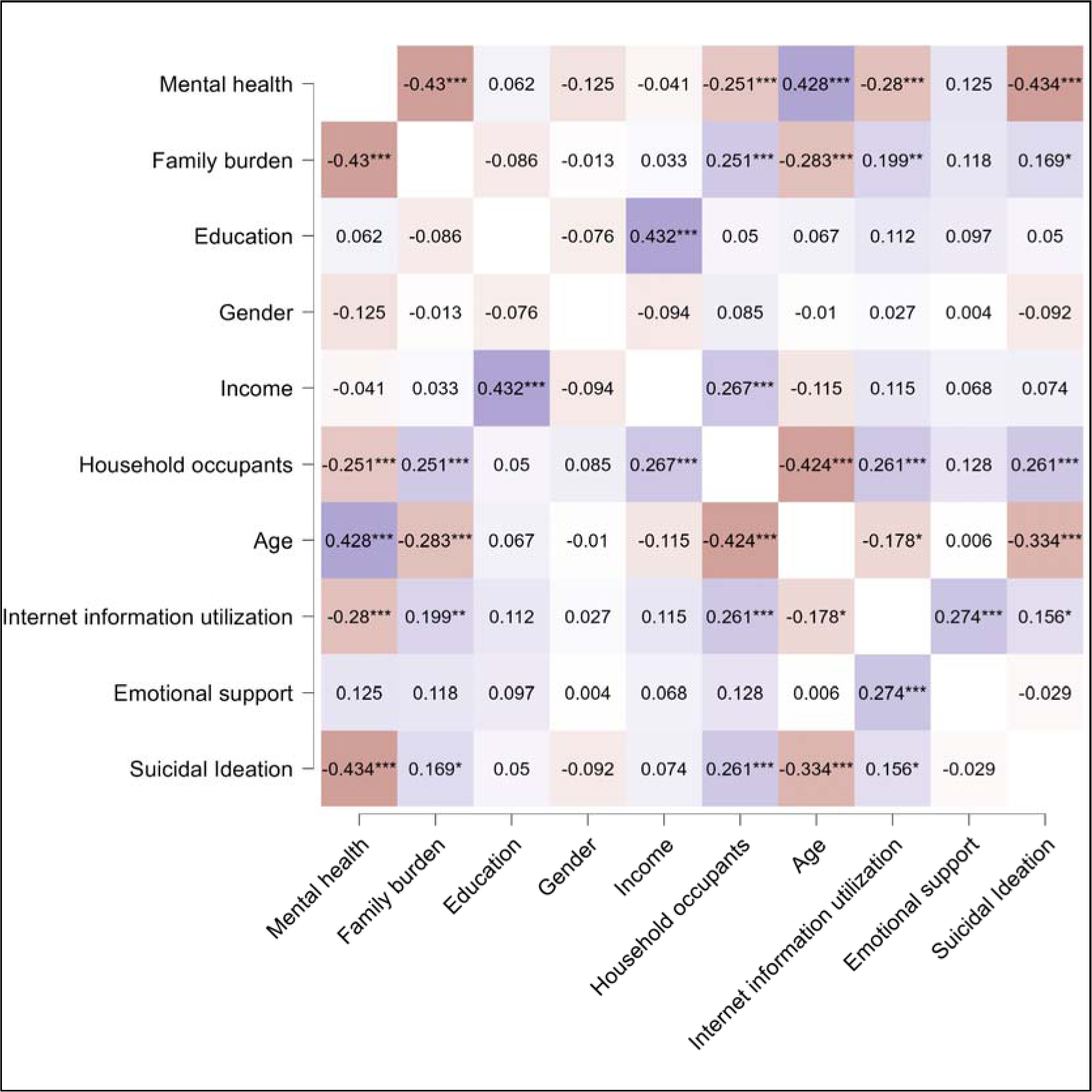
Correlation matrix of study variables.

The perceived burden on the family also appears to be a critical factor, as evidenced by its negative correlation with mental health (r = -0.430, *p < .001*), indicating that patients who feel their illness is a greater burden to their family may suffer more significant mental health challenges. This same perceived family burden showed a moderate positive correlation with suicidal ideation (r = 0.169), although this relationship was less pronounced. The number of household occupants was negatively correlated with mental health (r = -0.251), which may reflect the stress associated with larger household sizes, and this factor also showed a slight positive correlation with suicidal ideation (r = 0.118), suggesting a complex interaction between the patient’s immediate social environment and their mental well-being. Notably, age presented a robust negative correlation with suicidal ideation (r = -0.283), indicating that younger patients in this study were more prone to such thoughts compared to their older counterparts. This may reflect generational differences in dealing with illness or the accumulation of coping mechanisms over time.

Building upon the correlation findings, the subsequent linear analysis (Figure 8) refined our understanding of the factors influencing mental health and suicidal ideation, with a particular focus on the participants from Rural Appalachia compared to those from the rest of the United States. The analysis confirmed consistency across regions.

**Figure 8.**
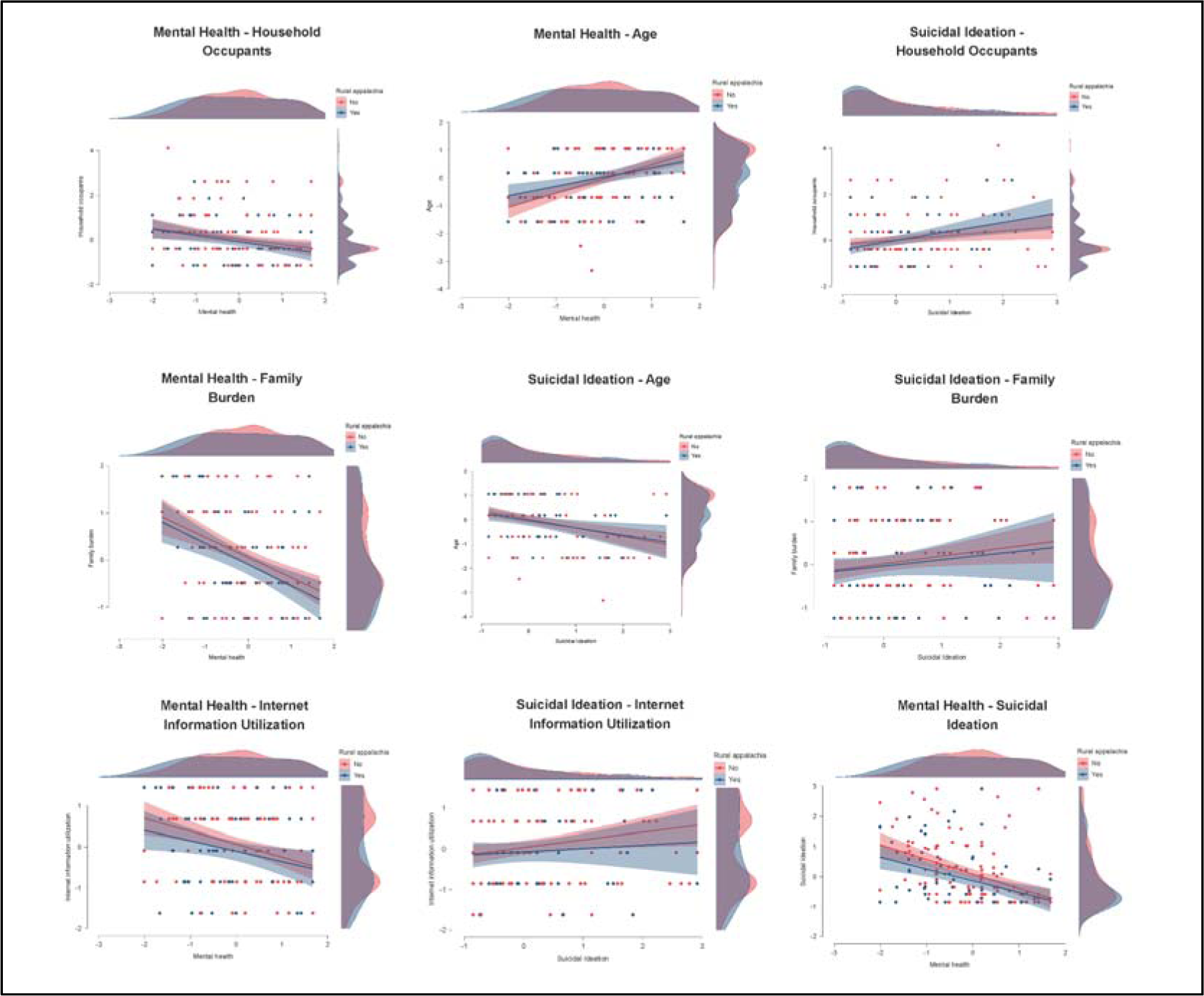
Linear relationships between mental health, suicidal ideation, internet information utilization, family burden, age, and household occupants across participants from Rural Appalachia and the rest of the US states.

### Qualitative findings from the survey

In our qualitative exploration of the experiences of cancer patients, several interrelated themes emerged, each illuminated by the participants’ poignant narratives. These themes, as noted in Table 2, included **(a)** the need for support, **(b)** the quality of care received, **(c)** the adequacy of information provided, and **(d)** the financial burdens faced during treatment. These findings offer a multifaceted perspective on the psychosocial challenges encountered by cancer patients and underscore the complexity of their mental health needs.

**Table 2.**
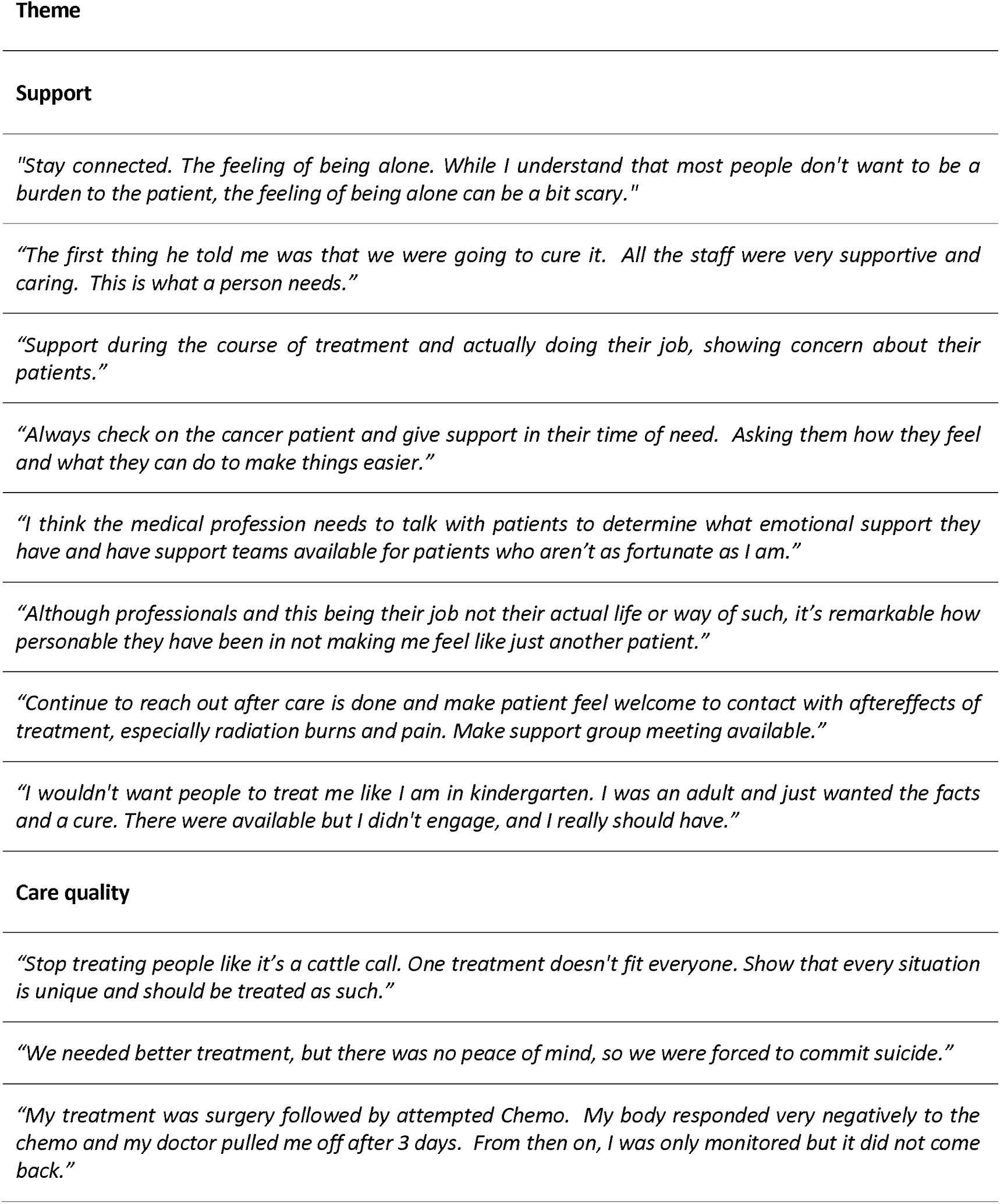
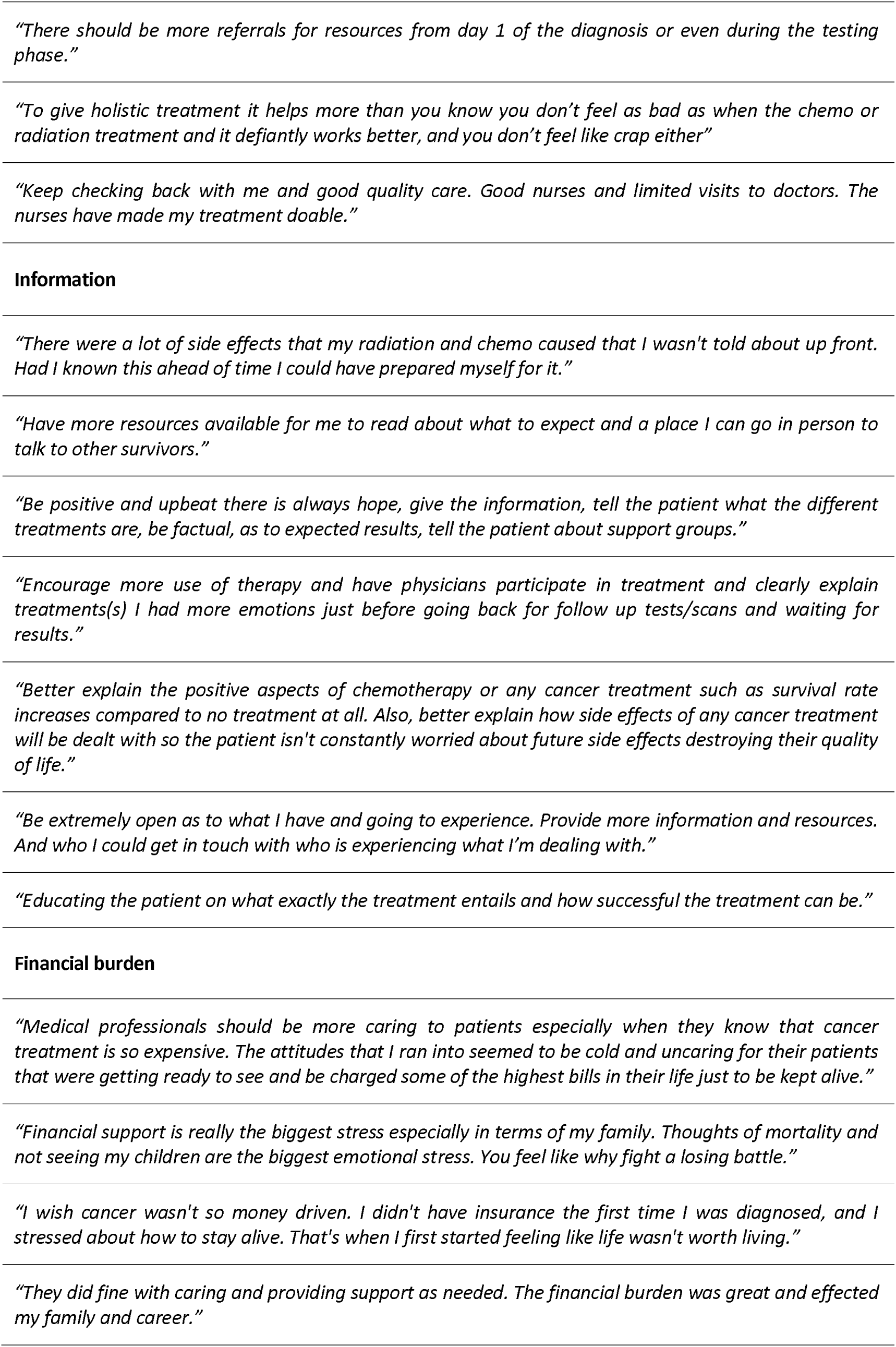
Sample quotes of study different participants.

#### Support

The critical role of social and emotional support was a recurring theme. One participant expressed, *“Stay connected. The feeling of being alone… the feeling of being alone can be a bit scary.”* This sentiment emphasizes the psychological impact of isolation and the value of a supportive presence. Another shared, *“Just being there for them, even if they say they don’t need anything or anyone… Just be there and recognize the signs.”* These narratives underscore the nuanced nature of the support required, which extends beyond mere physical presence to an empathetic understanding of the patient’s emotional state.

#### Care Quality

Personalized care emerged as a vital aspect, with participants advocating for treatment that recognizes individual needs. *“Stop treating people like it’s a cattle call. One treatment doesn’t fit everyone,”* one patient lamented, highlighting the need for more tailored care approaches. This theme was echoed in reflections on the balance between professional care and personal connection: *“Although professionals and this being their job not their actual life or way of such, it’s remarkable how personable they have been in not making me feel like just another patient.”*

#### Information

The importance of transparent and comprehensive information about treatment options, side effects, and outcomes was clearly articulated. A participant noted, *“There were a lot of side effects…that I wasn’t told about up front.”* This lack of preparation for treatment side effects points to a significant gap in patient education and communication, suggesting that more thorough and upfront information dissemination could significantly impact patient well-being.

#### Financial Burden

The financial strain associated with cancer treatment was frequently mentioned. One participant’s statement captures the essence of this burden: *“Financial support is really the biggest stress, especially in terms of my family.”* This theme underlines the mental toll of financial concerns, which compounds the stress of the illness and treatment, exacerbating mental health challenges.

### Qualitative findings from in-depth interview

#### Patient 1

The patient, a middle-aged man working from home, was unexpectedly diagnosed with colon cancer following a routine colonoscopy. In the initial stages of his cancer journey, the patient vividly recounts the jarring divergence between his expectations and the subsequent reality. His expression—“I was expecting good news, and it wasn’t”— poignantly captures the sharp pivot from anticipated health to the reality of illness, as seen in Figure 9.P1.C. This abrupt transition is marked by a psychological shock, a sentiment echoed in the analysis which shows emotional oscillations at the point of diagnosis. As the patient’s account unfolds, it sheds light on a healthcare system that tends to emphasize quick physical interventions, potentially at the expense of holistic patient support, a theme that is recurrent in his narrative. The proactive stance of his physician, as he recalls—“She doesn’t want to lose any of her patients… She said, well, let’s get it done”—can be seen as representative of a clinical ethos that favors rapid action, potentially neglecting the patient’s emotional and psychological needs during a critical time. The sentiment analysis in Figure 9.P1.D further corroborates the patient’s experience of emotional upheaval following his diagnosis. His account, “For several days? It really didn’t seem real… you think you’re invincible until you find out otherwise,” resonates with a common psychological pattern in the face of grave health news, where initial denial eventually succumbs to acceptance of reality. The absence of formal psychological support in his care underscores a significant lacuna in cancer care, as does his reliance on informal family support networks, highlighted in his remark—“It was mainly just talking to my wife and kids and stuff.” His reflections on the communication methods used by healthcare providers—“I don’t know if one is any better than the other”—bring to the fore the complexities involved in imparting sensitive health information. This sentiment, captured in the sentiment analysis, reflects the intricacies healthcare systems face in striking a balance between clinical efficiency and the need for empathetic patient engagement. The narrative also details the patient’s post-diagnosis treatment and care, including surgery and an abbreviated chemotherapy regimen, underscoring a disjunction between clinical treatment and sustained patient support, a gap evident in Figure 9.P1.D. His contemplation—“I had to go in, get just a follow-up visit… And then lots of and then the afterward treatment started a couple of months later”— signifies a disconnect in ongoing care coordination, pointing to the need for a more integrated approach in the continuum of cancer care.

**Figure 9.**
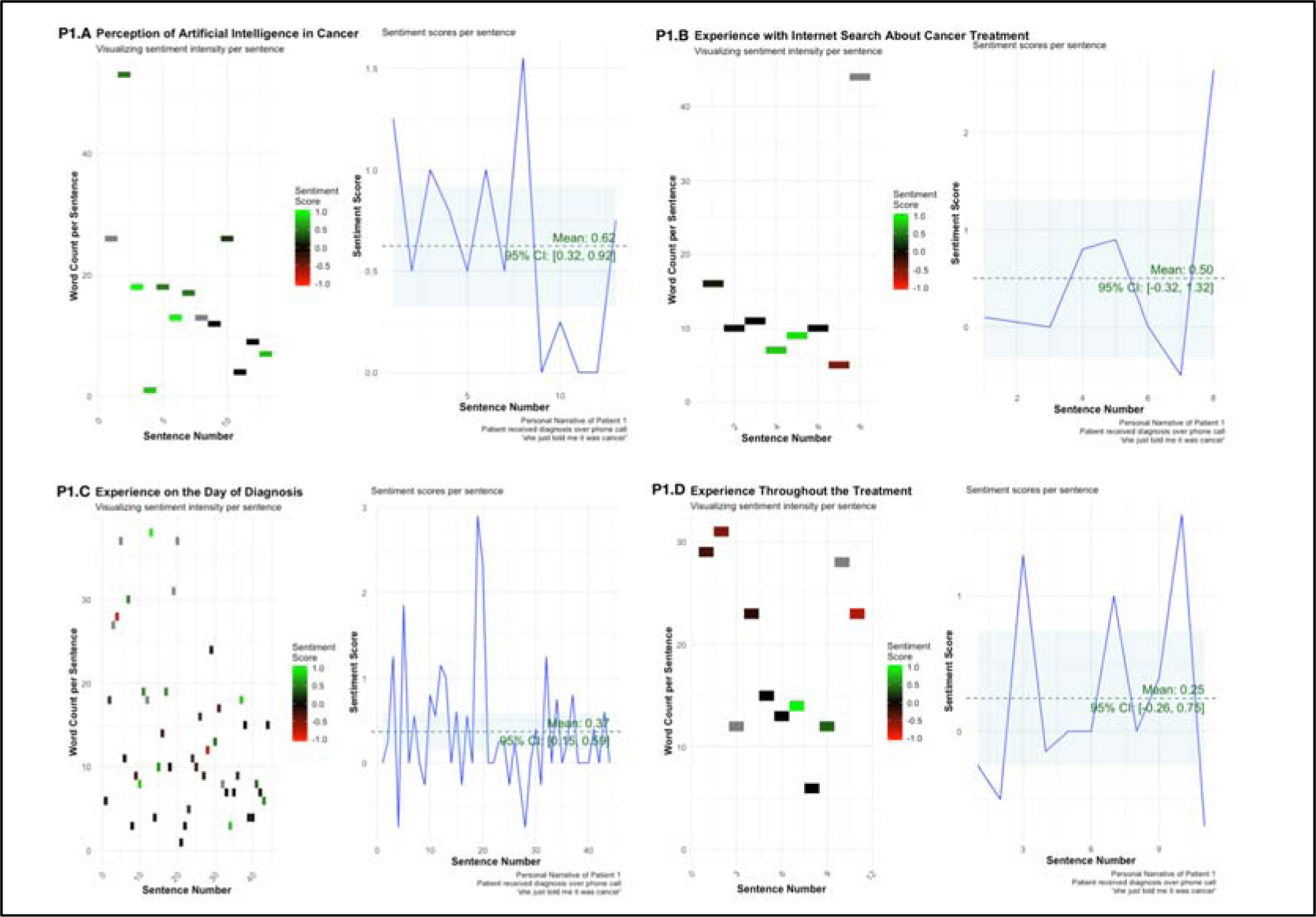
Patient 1 - Sentiment of Narratives Across Cancer Care Continuum. P1.A illustrates sentiment intensity per sentence in narratives discussing the perception of artificial intelligence (AI) in cancer, with a mean sentiment score highlighted. P1.B charts sentiment fluctuations in experiences with internet searches about cancer treatment, including a confidence interval around the mean sentiment. P1.C depicts sentiment on the day of diagnosis, showing a diverse range of emotional responses. P1.D visualizes sentiment throughout the treatment process, indicating overall sentiment trends and variance across the patient experience.

The patient’s experiences with conflicting online health information, encapsulated in his statement—“Some things will tell you, well, cancer feeds on sugar. And then other things would say, drink lots of carrot juice”—highlight the challenges and confusion that often accompany internet health searches, as visualized in Figure 9.P1.B. This underscores the pressing need for dependable, evidence-based information that could be augmented by AI, to navigate the intricate health landscape post-diagnosis (Figure 9.P1.A).

#### Patient 2

The patient was a 50 to 60 year-old man. His medical background is unique in that he was diagnosed with colorectal cancer, not through direct evidence in the colon or digestive tract, but through metastasis to the lungs, as indicated by markers—a nuanced detail that adds complexity to his care and emotional processing of the disease. On the day of diagnosis, the sentiment scores in Figure 10.P2.C suggest a stark oscillation of emotions, which the patient articulates as a mix of shock and disbelief. He describes an “odd way” of diagnosis, where a routine colonoscopy led to an incidental finding of lung spots during an X-ray. His reaction to the phone call from the pulmonologist—“I was kind of more stunned and just sort of shocked”—captures the unexpected and disorienting nature of his diagnosis. The sentiment analysis corroborates this narrative of sudden emotional upheaval.

**Figure 10.**
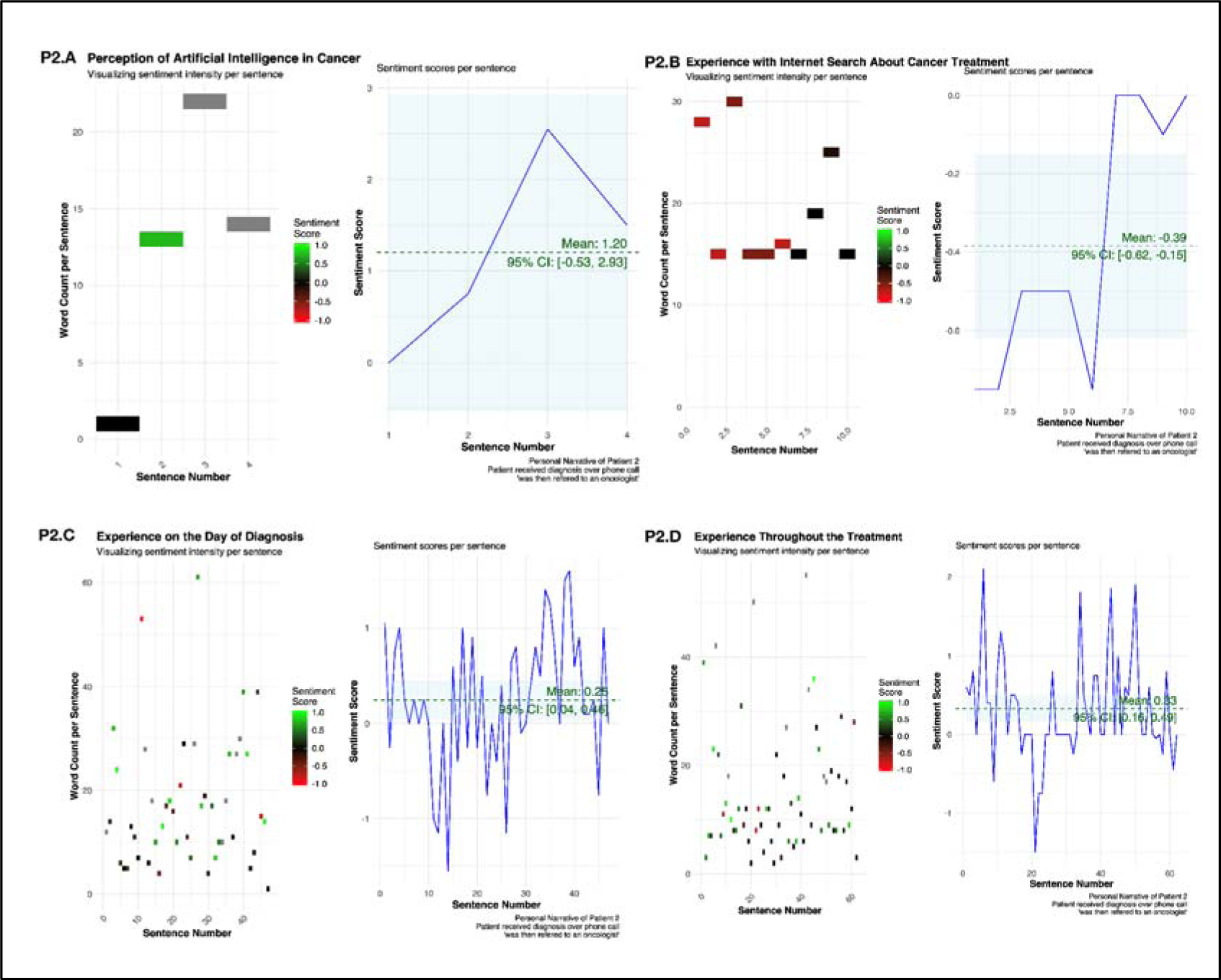
Patient 2 - Sentiment of Narratives Across Cancer Care Continuum. P2.A illustrates sentiment intensity per sentence in narratives discussing the perception of artificial intelligence (AI) in cancer, with a mean sentiment score highlighted. P2.B charts sentiment fluctuations in experiences with internet searches about cancer treatment, including a confidence interval around the mean sentiment. P2.C depicts sentiment on the day of diagnosis, showing a diverse range of emotional responses. P2.D visualizes sentiment throughout the treatment process, indicating overall sentiment trends and variance across the patient experience.

The patient’s journey is marked by three separate diagnoses over the years, each introducing its own set of treatments and emotional responses. Initially, surgical removal of tumors was successful, but subsequent recurrences led to chemotherapy and radiation, with the latter deemed ineffective due to the cancer’s resistance. These medical interventions and their impacts are depicted in the sentiment analysis of Figure 10.P2.D, where a relatively subdued positive mean sentiment belies the deeper emotional struggle conveyed by the patient’s reflections on his recurring battle with the disease. Mental health assistance was mentioned as part of the comprehensive care offered by his medical center, including a support group and the availability of oncology therapy, social workers, and dietitians. However, the patient points to the informal, empathetic interactions with nurses during his chemotherapy sessions as a significant source of emotional support—a sentiment not captured in clinical reports but reflected in the patient’s narrative and the sentiment analysis.

His experience with seeking information on the internet was fraught with confusion, leading to a negative sentiment score in Figure 10.P2.B. The patient acknowledges the anxiety and misinformation that can arise from unguided online searches, which reinforces the need for patient-centric, reliable, and comprehensible medical information. Lastly, the patient expresses a positive perception of AI, as shown in the sentiment analysis of Figure 10.P2.A. He sees value in technology that can simplify complex medical information, provide insights into available treatments, and facilitate participation in clinical studies. His proactive approach to understanding his condition and treatment options through AI reflects a desire for autonomy and empowerment in his care journey.

#### Patient 3

The patient, senior citizen, details a protracted 20-year journey with cancer that began with a prostate cancer diagnosis in early 2000s. His treatment for prostate cancer led to radiation proctitis and subsequent significant internal bleeding, which was controlled for a time with a formaldehyde-related procedure. It was during a routine checkup for prostate cancer, which had been in control, that his hemoglobin levels indicated life-threatening internal bleeding, prompting further investigation. This investigation uncovered a lump on his throat, which was biopsied and diagnosed as stage four squamous cell carcinoma of the head and neck, metastasized and aggressive.

The day he learned of his GI diagnosis was marked by a lack of communication and oversight from his healthcare providers, as illustrated by the sentiment analysis in Figure 11.P3.C. The sentiment intensity here is notably volatile, reflecting the tumultuous emotional response to his diagnosis. His experience was complicated when, after numerous discussions with a panel of doctors about his head and neck cancer, a cancer research scientist, identified colon cancer in the scans—a significant oversight by his medical team. Throughout his journey, the patient describes a dearth of mental health support from his initial team of doctors, who, after a fraught interaction, were not consulted again: “Not the original group of doctors… We have never gone back.” Instead, he found strength and resilience within himself, remarking, “I’m a pretty tough old bird,” and in the planning and preparation for his family’s future in the face of potential mortality.

**Figure 11.**
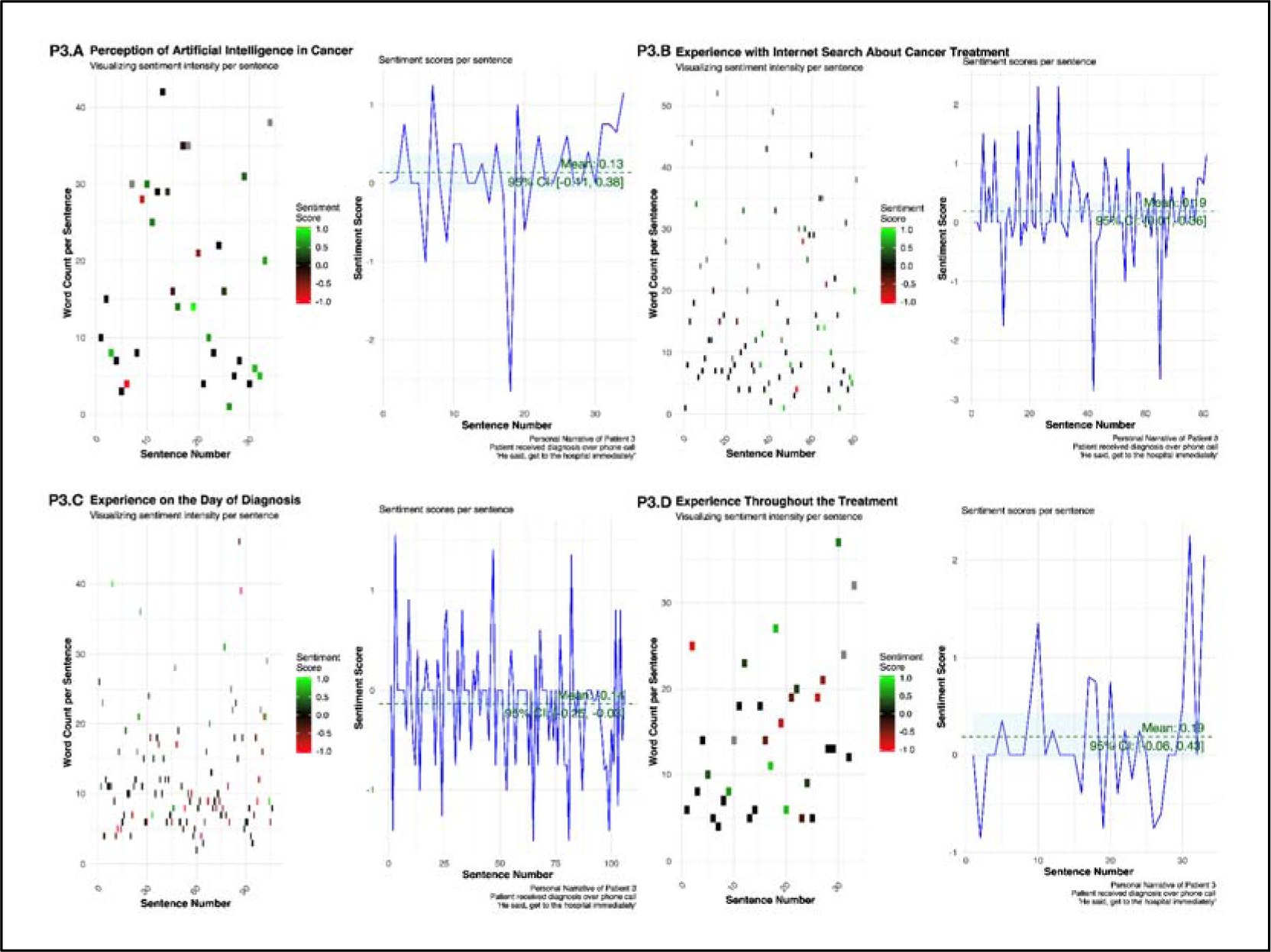
Patient 3 - Sentiment of Narratives Across Cancer Care Continuum. P3.A illustrates sentiment intensity per sentence in narratives discussing the perception of artificial intelligence (AI) in cancer, with a mean sentiment score highlighted. P3.B charts sentiment fluctuations in experiences with internet searches about cancer treatment, including a confidence interval around the mean sentiment. P3.C depicts sentiment on the day of diagnosis, showing a diverse range of emotional responses. P3.D visualizes sentiment throughout the treatment process, indicating overall sentiment trends and variance across the patient experience.

His experience with the internet in seeking information was characterized by discernment and selectivity. Despite the sentiment analysis in Figure 11.P3.B suggesting a neutral average sentiment, he recounts his cautious approach to online resources: “We’re college educated Americans, and we’ve been the route, and we know how to read and read reviews and filter out what’s good, what’s bad.” The patient holds a positive view of AI, as indicated by the favorable sentiment scores in Figure 11.P3.A. He articulates the potential value of AI in managing a cancer diagnosis: “Absolutely… that’s something that has to come to be, and it will allay a lot of the fears, anxiety, depression, concern of a cancer patient.” His advocacy for accessible and understandable information via AI aligns with his proactive stance on patient education and empowerment.

#### Patient 4

The patient, senior citizen, faced multiple cancer diagnoses and treatments over several years, with a significant medical background involving Hodgkin’s disease, sarcoma, and colon cancer, also had a complex family situation with a history of cancer affecting close family members. On the day of his diagnosis, the patient described a visit to the clinic with his wife, where he received confirmation of Hodgkin’s disease from his Hematology Oncologist: “My primary care physician at the time said, I think you may have Hodgkin’s disease… My wife at the time went with me. We spent about an hour talking with him, and he told me what his diagnosis was.” The sentiment scores on Figure 12. P4.A during this recount show a moderate positive peak, suggesting a moment of clarity or acceptance amid the diagnosis narrative. The gap between diagnosis and treatment initiation was about ten days, a time described by the patient as active and decisive, involving family discussions and personal research: “I got my two kids, we got them on a conference call, discussed with them what was found, what the treatment was and what the process would be.” Figure P4.B shows the sentiment during this period as fluctuating, indicating the patient’s varying emotional responses during the anticipatory phase of treatment.

**Figure 12.**
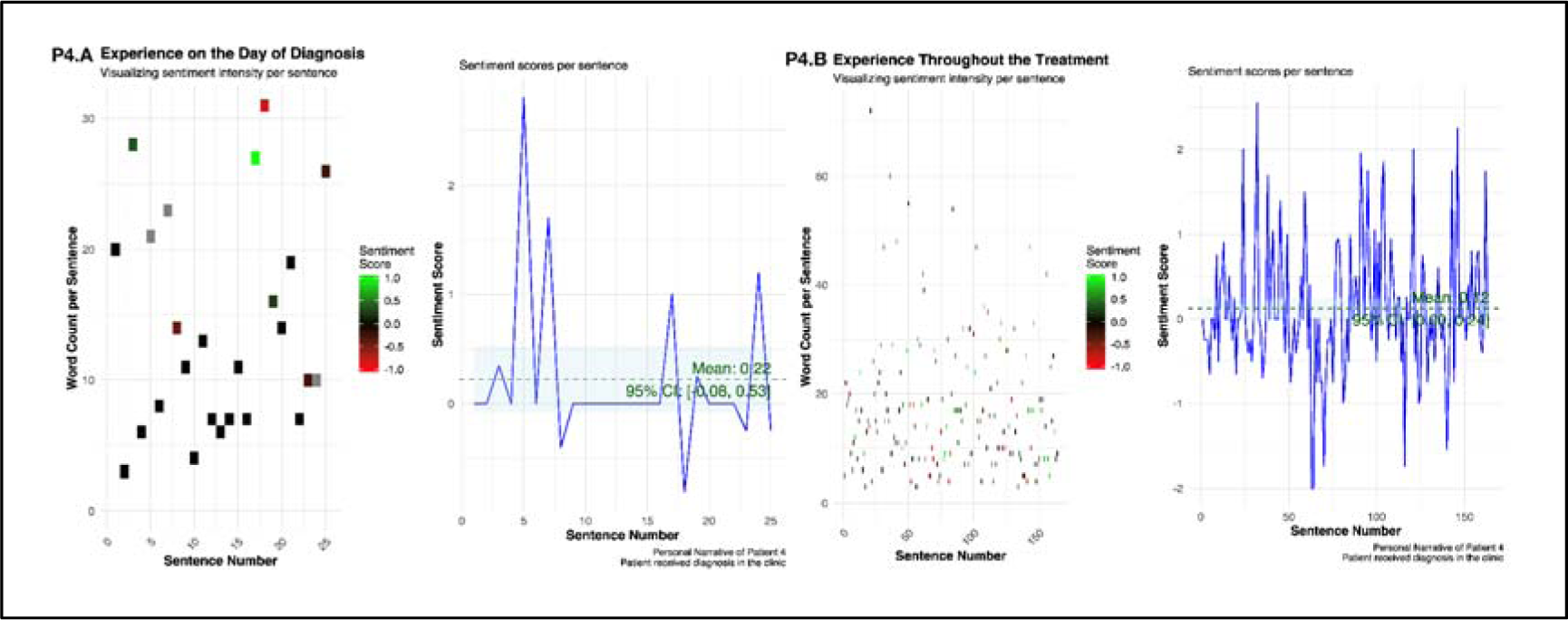
Patient 4 - Sentiment of Narratives Across Cancer Care Continuum. P4.A depicts sentiment on the day of diagnosis, showing a diverse range of emotional responses. P4.B visualizes sentiment throughout the treatment process, indicating overall sentiment trends and variance across the patient experience. This patient did not use Internet or had any opinion on artificial intelligence in cancer care.

Mental health assistance seemed to be self-managed initially, with the patient opting not to seek help despite the available services: “I was sort of stoic about it… I didn’t have a real good feel for mental health at the time, which was a mistake.” This sentiment reflects the low sentiment scores observed in Figure12. P4.B, representing his initial reluctance to engage with mental health services. His treatment experience was communicated via regular emails with his doctor, a method that suited his preference and allowed him to maintain control and awareness of his treatment: “Fortunately, my doctor was very email responsive, so I kept in touch with him…” This communication style likely contributed to the positive sentiment scores seen in Figure 12 P4.B, as the patient felt supported and informed.

## Discussion

### Discussion of Quantitative Findings

In the exploration of suicidal ideation among GI cancer patients, a significant distinction emerges between passive and active forms of such thoughts. Passive ideation, characterized by a desire not to wake up or a wish to be dead, is observed to be more prevalent than active plans for suicide. This trend aligns with psychiatric research, which has consistently shown that the transition from passive to active ideation is a critical juncture where the risk of suicide attempts escalates [36,37]. For example, Wastler et al. in 2022 demonstrated that both passive and active ideation are associated with increased odds of attempting suicide [38]. Additionally, May et al. in 2015 highlighted that passive ideates are at significant risk for a future attempt, further emphasizing the importance of recognizing and addressing passive suicidal ideation [39]. The data also reveals a notable decrease in the intensity of suicidal thoughts as they become more action-oriented, suggesting a critical window for intervention. This presents an opportunity for healthcare providers to implement strategies aimed at identifying and supporting patients with passive ideation, thereby potentially disrupting the progression to more severe forms of suicidal thoughts.

Furthermore, the emotional challenges faced by cancer patients during their treatment journey highlight a complex and often counterintuitive emotional landscape. The most distressing phase was frequently found to be the post-treatment recovery period, surpassing the initial diagnosis in terms of mental and emotional strain. This suggests that the end of active treatment does not necessarily correspond with a decrease in psychological distress, possibly due to feelings of vulnerability or anxiety that arise when regular medical support diminishes. Such insights call for a reassessment of post-treatment care models to provide more comprehensive support during this critical phase. In 2022 Moura et al. also emphasized the need for early-stage post-cancer treatment care outside of regional cancer centers [40]. Therefore, our research advocates for a holistic and continuous model of care, integrating mental health as a fundamental aspect (not an optional extension) of cancer treatment and recovery, thereby ensuring comprehensive support for patients throughout their journey.

The finding that the post-treatment recovery period is frequently the most distressing phase, surpassing the initial diagnosis in terms of mental and emotional strain, is supported by Costanzo et al., who found that the adjustment to life after treatment for breast cancer can be distressing, despite the expectation of relief [41]. In 2019, a study by Trevino et al. highlighted the distress experienced by cancer survivors [42]. Additionally, in 2016, Borowski et al. discussed the influence of individual, family, and community-level factors on the mental health of survivors of colorectal cancer, indicating the multifaceted nature of distress in cancer survivors [43].

The correlation analysis offers crucial insights into how psychosocial factors affect cancer patients, highlighting two notable correlations with direct implications for mental health interventions. There is a nuanced relationship where internet information utilization correlates negatively with both mental health and suicidal ideation. This duality suggests that while seeking information online can be a coping mechanism that potentially diminishes suicidal ideation by reducing uncertainty and fear, it can simultaneously be linked to poorer mental health outcomes. This paradox could stem from various causes: individuals grappling with mental health challenges might turn to the internet for help, yet the information they find could exacerbate their anxiety, especially if it is complex, overwhelming, or grim.

Contrary to expectations, the empowering effect of accessible information does not always translate into improved mental health. Instead, this correlation paints a picture where the quality and presentation of information are critical. Patients may suffer increased distress if the health information they encounter online is not well-curated to their needs. This highlights the imperative for healthcare providers to guide patients toward information that is not only reliable and relevant but also presented in a way that is comprehensible and reassuring. By doing so, the internet’s vast resources can be harnessed to support mental health effectively, ensuring that patients benefit from online information without incurring additional psychological distress.

Our findings that the empowering effect of accessible information does not always translate into improved outcome (mental health) is supported by Diviani et al., who in 2015 found that low health literacy plays a role in the evaluation of online health information, indicating that individuals with lower health literacy may struggle to assess the quality of the information they encounter [44]. In 2021, Newman et al. suggested that the quality and accessibility of information are crucial for promoting mental health and well-being [45]. In the same line, Wong & Cheung in 2019 acknowledged the importance of understanding patients’ behaviors and needs along with the quality and presentation of information [46].

### Discussion of Qualitative Findings from the Survey

The open-ended survey responses of GI cancer patients feature the critical role of social and emotional support in their cancer journey. The finding resonates with the existing literature in psycho-oncology, which emphasizes the protective role of social support against psychological distress in cancer patients [47,48]. Additionally, the majority of patients with gastrointestinal (GI) malignancies are older, highlighting the importance of understanding the unique support needs of this patient population [49]. Evidence also suggests that perceived social support may be more critical than actual support available or received in terms of quality of life outcomes [50]. Studies have also demonstrated that emotional support positively impacts mental health by decreasing a sense of loneliness in individual advanced GI cancer patients and caregivers [51].

Patients’ perceptions of care quality and the desire for personalized treatment approaches highlight significant aspects of patient-centered care. The emphasis on individualized treatment resonates with the principles of person-centered therapy [52], suggesting that recognizing and addressing individual patient needs contributes significantly to treatment efficacy and patient satisfaction. The principles of person-centered therapy emphasize the importance of understanding and addressing individual patient needs [52]. In 2018, Loonen et al. discussed the significance of person-centered care in cancer survivorship, aiming to empower survivors and support self-management [53]. In 2020 Crabtree et al. reported the importance of recognizing individual needs in cancer survivorship [54]. Thus, our study advocates for a balance between clinical efficiency and personalized care, as the de-personalization of treatment can negatively impact patient morale and trust in the healthcare system.

The significance of transparent and comprehensive information provision, as highlighted by the participants, reflects the concept of shared decision-making in healthcare. Patients’ desire for thorough information about treatment options, side effects, and outcomes points to the importance of informed consent and patient autonomy. The financial strain associated with cancer treatment also emerged as a profound stressor in our cohort. Existing studies have also associated financial strain with increased psychological symptoms in cancer patients, including anxiety and depression [55]. Additionally, the financial impact of cancer has been identified as a stressor that can precipitate posttraumatic stress disorder [56]. Patients’ concerns about financial burdens reflect the broader socio-economic context of healthcare, where studies have acknowledged relationships between lower socio-economic position and reduced chances of early stage cancer diagnosis, impacting treatment outcomes and survival [57].

Although previously documented in the literature and acknowledged by healthcare authorities and practitioners, these concerns persistently emerged in our cohort. The recurring nature of these issues suggests that they still need to be addressed in the current healthcare framework. Despite the awareness of these challenges within the medical community, there appears to be a gap between understanding these concerns and implementing practical solutions. This underscores an urgent need for further research and targeted intervention by medical experts and policymakers to enhance the patient experience and address these enduring concerns.

### Discussion of Qualitative Findings from In-Depth Interviews

The in-depth narratives of the four cancer patients in this study present a comprehensive picture of the psychological and emotional landscape of individuals undergoing cancer treatment. These narratives, while unique in their details, collectively highlight several critical themes in the realm of cancer care: the psychological adaptation to cancer diagnosis, the importance of support systems and provider-patient relationship, and the emerging significance of AI and digital tools in healthcare.

#### Psychological Adaptation to Cancer Diagnosis

The journey from shock and denial to eventual acceptance, as illustrated by patient 1, is a common psychological pathway for many cancer patients. This adaptation process is often complex and multidimensional, necessitating structured psychological support, including pre-habilitation assessment before treatment, rehabilitation assessment post-treatment, and health promotion assessment at the end of treatment [58]. The narratives of our cohort reveal, there exists a notable gap in formal psychological care within the healthcare system. Despite the availability of evidence-based clinical practice guidelines for psychosocial care of cancer patients, there are challenges in the utilization of professional psychological care, indicating the need for improved implementation and accessibility of such services [59]. Patients in our study relied heavily on informal support networks, such as family and friends, highlighting a critical area for improvement in providing comprehensive cancer care.

#### Importance of Support Systems and Provider-Patient Relationships

The experiences of patients 2 and 4, in particular, shed light on the varied forms of support systems that cancer patients lean on. Patient 2’s story highlights the emotional support garnered from empathetic interactions with nurses, suggesting that these relationships play a crucial role in patient well-being [60,61]. Patient 4’s narrative illustrates the complexities of managing cancer within a family context and the challenges of self-managed mental health, underscoring the need for integrated care systems that include mental health services [62]. The narratives underline the significance of empathetic care in the healthcare setting. Patients often remember and value the compassionate interactions they have with their caregivers, more than the clinical aspects of their care. This empathetic approach is not only beneficial for emotional well-being but can also positively impact treatment outcomes [62]. The narratives also collectively emphasize the necessity of effective communication and holistic care in cancer treatment. Patient 3’s two-decade-long battle with cancer, marked by periods of ineffective communication with healthcare providers, highlights the critical need for clear and consistent communication and oversight in chronic disease management [63,64]. Holistic care, addressing both the physical and psychological needs of patients, emerges as a key element in improving the overall quality of cancer care.

#### Role of AI and Digital Tools in Cancer Care

Patients 2 and 3 bring attention to the growing trend of utilizing AI and internet resources in navigating their health journey. Their proactive engagement with digital tools reflects a broader shift towards patient empowerment and self-advocacy in healthcare. This trend underscores the importance of reliable, easily accessible medical information and AI-augmented tools to guide patients through the complexities of cancer care, enabling more personalized and informed decision-making [65,66].

### Limitations

In addressing the limitations of our study, it is essential to consider several factors. The sampling method, primarily based on self-selection via a web-based survey distributed through social media and open cancer forums, may lead to selection bias and limit the generalizability of our findings to the broader population of individuals with GI cancer in the US. Furthermore, the reliance on self-reported data raises concerns regarding the accuracy and potential bias in participant responses, although a control question was included to enhance response integrity. The limited number of follow-up interviews, restricted to just four individuals, may not sufficiently represent the diverse experiences of the targeted population, thus potentially narrowing the scope of qualitative insights. The exclusive use of online platforms for data collection could inadvertently exclude individuals lacking internet access or digital literacy.

## Conclusion

Our study offers a comprehensive exploration of the psychosocial experiences of gastrointestinal cancer patients, providing valuable insights into their mental health challenges, coping mechanisms, and perceptions of healthcare communication and artificial intelligence in cancer care. Key findings revealed a notable prevalence of suicidal ideation, particularly in the more passive forms. However, the intensity of such thoughts decreased as they transitioned towards concrete planning, highlighting critical intervention points to prevent progression to more severe stages. The most significant mental distress was reported not at the time of diagnosis, as might be expected, but during the post-treatment recovery phase. This finding underscores the necessity for continuous psychological support extending beyond the completion of medical treatment. The study also illuminated the critical role of psychosocial factors, such as family burden and emotional support, in influencing the mental health of GI cancer patients. Patients who perceived their illness as a substantial burden to their families experienced greater mental distress, emphasizing the importance of holistic care that includes family counseling and support services.

Furthermore, the role of artificial intelligence emerged as a promising avenue for enhancing patient understanding of their condition and treatment options. The positive perception of AI by patients suggests its potential to simplify complex medical information and empower patients in their care journey. This study also highlighted the need for effective communication strategies in healthcare. Patients valued clear, empathetic communication and the provision of comprehensive, understandable information from their healthcare providers.

## Author Contributions

Conceptualization, A.C.; methodology, A.C.; validation, A.C. I.A., M.A.; formal analysis, A.C. and Y.S.; investigation, Y.S. and A.C.; resources, A.C.; data curation, A.C.; writing—original draft preparation, A.C., I.A., M.A., and S.E.; writing—review and editing, A.C., I.A., and S.E..; visualization, A.C.; supervision, A.C.; project administration, A.C. All authors have read and agreed to the published version of the manuscript.

## Data Availability

The datasets analyzed during the current study are not publicly available as we do not have permission to share the interview data.

## Ethics approval

This study received ethical approval from the West Virginia University Institutional Review Board under protocol number 2212691613. The study qualified for the WVU Flexibility Review Model, as it involves minimal risk and adheres to the Belmont Report’s ethical principles. Approval was granted on February 7, 2023.

## Consent to participate

Informed consent was obtained from all individual participants included in the study.

## Data Availability

All data produced in the present study are available upon reasonable request to the authors

